# Genetic Interplay Between White Matter Hyperintensities and Alzheimer’s Disease: A Brain-Body Perspective

**DOI:** 10.1101/2024.09.27.24314431

**Authors:** Manpreet Singh, Kimia Shafighi, Flavie E. Detcheverry, Fanta Dabo, Ikrame Housni, Sridar Narayanan, Sarah A. Gagliano Taliun, Danilo Bzdok, AmanPreet Badhwar

## Abstract

MRI-detected white matter hyperintensities (WMH) are often recognized as markers of cerebrovascular abnormalities and an index of vascular brain injury, and are frequently present in individuals with Alzheimer’s disease (AD). Given the emerging bidirectional communication between the brain-body axis in both WMHs and AD, it is important to understand their genetic underpinnings across the whole body. However, literature on this is scarce.

We investigated the brain-body axis by breaking down heritability estimates of these phenotypes across the whole body, – i.e., partitioning heritability. Our aims were to identify genetic underpinnings specific to WMHs, and common between WMHs and AD, by assessing (a) the partitioned heritability of WMHs and AD across the brain-body axis with tissue-specific annotations, (b) the partitioned heritability of WMHs and AD across the brain-body axis with cell-specific annotations, and (c) the genes associated with WMHs and AD, and verifying their expression levels across the whole body.

Our tissue-specific analysis revealed that WMH-associated SNPs were significantly enriched in tissues beyond the brain, namely liver, cardiovascular, and kidney – with liver being a common tissue enriched for both WMHs and AD. Our cell-specific analysis showed enrichment of vascular endothelial cells across the tissue types enriched for WMHs, highlighting their central role in the development of WMHs. Additionally, our gene-level analysis highlighted overlapping patterns of tissue enrichment for both WMHs and AD, and showed interactions between WMH and AD associated genes.

Our findings provide new insights into the systemic influences potentially contributing to WMH pathology, in particular, multi-system endothelial disorder. We hope that our multisystemic genetic findings will stimulate future WMH-research into specific pathways across the brain-body axis.

## 1. INTRODUCTION

Established magnetic resonance imaging (MRI) markers of vascular brain injury include white matter hyperintensities (WMHs), silent brain infarcts, microbleeds and enlarged perivascular spaces ^1^. A meta-analysis by Debette et al. ^1^ found that among these markers, extensive WMH volume was significantly associated with an increased risk of dementia, including Alzheimer’s disease (AD) – the most prevalent of the dementias ^2^. WMHs appear as bright spots on on T2-weighted fluid attenuated inversion recovery (FLAIR) MRI ^3^, and often represent areas of axon injury/loss, demyelination, and some degree of edema ^4,5^. Suggested vascular pathogenic mechanisms of WMHs include endothelial and associated blood-brain barrier dysfunction, hypoperfusion due to altered cerebrovascular autoregulation and reactivity, and cerebral amyloid angiopathy ^6–8^. The prevalence of WMHs increases significantly with advancing age ^9^, affecting 11-21% of people around age 64 and rising to 94% by age 82 ^4,9^. While observed in cognitively healthy adults, WMHs are more frequently present in individuals with AD ^10–12^. Moreover, a growing body of evidence shows a link between their presence and cognitive impairment, a key feature of AD ^1,13^.

WMHs are heritable, with family and twin studies estimating a genetic contribution of 55-73% ^14–17^. Heritability is the proportion of the total phenotypic variance in a trait that is attributable to the additive effects of common genetic variants, e.g., single nucleotide polymorphisms (SNPs) ^16^. To date, three genome wide association studies (GWAS) ^18–20^ and two GWAS-meta analyses ^21,22^ focusing on WMHs have identified multiple loci associated with WMH burden. The seminal study by Fornage et al. ^18^ identified one locus on chromosome 17q25, encompassing genes *WBP2, TRIM47*, *TRIM65*, *MRPL38*, *FBF1*, and *ACOX1.* Novel associations were revealed at *PLEKHG1* ^19^, along with confirmations of previous findings on *TRIM47* and *EFEMP1*, as shown by ^18^ and ^21^, respectively. Since then, additional studies using larger sample sizes (∼18,000 ^20^, ∼50,000 ^22^) have identified 18 additional loci for WMHs. Late-onset AD demonstrates similar heritability: 58-79% ^23,24^ and evidence suggests a potential genetic overlap between WMHs and AD, particularly involving inflammation-related genes ^25^. This overlap points to certain shared pathways that may contribute to the development of both phenotypes.

Moreover, WMHs are strongly associated with vascular risk factors like diabetes, smoking, and hypertension, all of which have widespread impacts across the body ^4,26^. While traditionally seen as a cerebral issue, recent research suggests that the presence of WMHs in the brain may reflect a broader multi-systemic condition. For example, (a) individuals with small vessel disease, in whom WMHs are a hallmark feature ^3^, show an increased risk of renal impairment ^27^, and (b) non-alcoholic fatty liver disease has also been associated with the presence of WMHs ^28^. Similarly, AD, though primarily considered a brain disorder, has also been linked to peripheral systems. In particular, the liver is involved in the clearance of amyloid-beta, a hallmark of AD pathology ^29^ and studies also indicate the involvement of systemic immune responses in AD ^29,30^.

Given the bidirectional communication between the brain-body axis in both WMHs and AD, it is important to understand the commonalities and differences of their genetic underpinnings across body systems. However, literature on this is scarce. One way to investigate the brain-body axis is by breaking down heritability estimates of these phenotypes across the whole body – i.e., partitioning heritability ^31^. Therefore, our aims are to identify genetic underpinnings specific to WMHs, and common between WMHs and AD, by assessing (a) the partitioned heritability of WMHs and AD across the brain-body axis with tissue-specific annotations, (b) the partitioned heritability of WMHs and AD across the brain-body axis with cell-specific annotations, and (c) the genes associated with WMHs and AD, and verifying their expression levels across the whole body.

## 2. METHODS

### 2.1 Genome Wide Association Studies With Publicly Available Summary Statistics

We used the GWAS catalogue (https://www.ebi.ac.uk/gwas/) to identify GWAS summary statistics for WMHs and AD. This catalogue provides comprehensive GWAS data on various traits. Our search in GWAS catalogue was conducted using the following keywords: “White matter hyperintensities”, “Alzheimer’s disease”. In addition we conducted a literature review to identify GWAS summary statistics for WMHs by using the PubMed scientific database with the following keyword combinations: “WMHs” AND “GWAS”; “White matter hyperintensities” AND “GWAS”; "White matter hyperintensities" AND "Genome wide association study". This search was completed in August 2024 and yielded 54 articles, from which we included only those published in English and for which summary statistics were available in our subsequent analysis (N=3). Summary statistics for AD were referenced from the GWAS catalogue (https://www.ebi.ac.uk/gwas/) (Supplementary Table S1).

### 2.2 Linkage Disequilibrium Score Regression (LDSC)

We employed linkage disequilibrium (LD) score regression (LDSC, version 1.0, https://github.com/bulik/ldsc) ^31^ to estimate the proportion of phenotypic variance attributable to common autosomal variants, expressed as SNP-based heritability (h^2^_SNP_), for WMH and AD studies. LD refers to non-random association of alleles at different loci in a population ^32^. LD score regression (LDSC) computes an LD score by summarising the correlations of a given SNP with all neighbouring SNPs within its downstream and upstream 100 kb flanks ^31^. The GWAS test statistic χ2 is then regressed against the LD score, from which the slope is rescaled to provide an estimate of h^2^_SNP_, attributable to all SNPs considered by the LD score analysis ^31^. For our analyses, we used precomputed LD scores based on 1,000 Genomes Project data focusing on the European ancestry population ^33^.

### 2.3 Partitioned SNP-heritability analysis

We conducted stratified LDSC (s-LDSC; version 1.0) to identify which tissue types are enriched for variants that significantly contribute to per-SNP heritability (h^2^_SNP_) of WMHs and AD. This analysis aimed to explain how specific genomic annotations (i.e., tissue-specific and cell-specific) disproportionately influence h^2^_SNP_. In this context, enrichment is defined as the ratio of heritability attributed to SNPs (h^2^_SNP_) within a given tissue type compared to the proportion of SNPs annotated for that same tissue type across the autosomes. We utilised pre-computed LD scores from the 1,000 Genomes Project European (EUR) subpopulation as the LD reference panel ^34^. These scores are based on common autosomal SNPs from the HapMap project, with the major histocompatibility complex (MHC) region excluded due to its dense gene content and long-range LD ^34^.

For data preparation, we utilised the LDSC script “munge_sumstats.py” which was run on summary statistics files (GRCh38) for WMH and AD studies. The LD weights were derived from non-MHC HapMap3 SNPs and applied in the regression analyses as ’weights_hm3_no_hla’. Our LD score regression analyses employed a full baseline model (baselineLD_v1.2) along with detailed annotations as discussed below, using the ’--overlap-annot’ parameter alongside minor allele frequency files (’1000G_Phase3_frq’).

#### 2.3.1 Tissue-specific heritability

To characterise tissue-specific partitioned heritability across human tissues, we performed s-LDSC on published GWAS summary statistics. This included data from three WMH studies and six AD studies, analysing phenotypes of WMHs and AD across ten published tissue-specific binary annotations ^31^. For each GWAS, enrichment of phenotype-associated SNPs within each tissue-type was calculated. The top three -log10(enrichment_p) values (representing strength of association) were used to interpret the data qualitatively and check for enrichment patterns across the GWAS studies. Thereafter, a 5% false discovery rate threshold was applied on enrichment_p to correct for multiple testing comparisons (enrichment_p < 0.05) to statistically ascertain tissue types enriched for each study following the qualitative assessment.

#### 2.3.2 Cell-specific heritability

To characterise the cell-specific contribution towards heritability of these phenotypes (WMHs, AD) within these enriched cell types, we partitioned heritability of phenotypes across published cell-specific continuous annotations ^35^ for all cell-types associated with tissues enriched for WMHs. -log10(enrichment_p) values were used to interpret the data qualitatively and check for enrichment patterns across the GWAS studies. A 5% false discovery rate threshold was applied to correct for multiple comparisons (enrichment_p < 0.05) to statistically identify cell types enriched across all WMH and AD studies after the qualitative analysis.

### 2.4 Gene-based analysis

#### MAGMA analysis

MAGMA was used to provide a gene-level resolution to complement the broader tissue-and cell-specific heritability insights from LDSC. We carried out a gene analysis (testing the joint association of variants) in MAGMA (version 1.10) ^36^ to identify genes associated with our two phenotypes, namely WMHs and AD. MAGMA’s gene analysis uses a multiple linear principal components regression to map all SNPs from inputted summary statistics to their respective genes, evaluating the collective impact of all variants within each of the protein-coding genes from the NCBI 38 database ^36^. This method leverages an F-test to determine gene *p*-values, which are computed efficiently compared to permutation-based methods. Gene-level association statistics were obtained by combining *p-*values within defined windows surrounding each gene – 35 kb upstream and 10 kb downstream – accounting for linkage disequilibrium using data from the European super-population of the 1,000 Genomes Project Phase 3 ^37^. Significance was defined by Bonferroni-corrected *p*-values (*p* < 0.05/total number of protein-coding genes).

#### Characterization of MAGMA-prioritised genes

Statistically significant genes (*p* < 0.05/number of protein coding genes) identified for WMHs and AD were then assessed and compared for their enrichment in different tissues and cell types across the brain-body axis using the human protein atlas (https://www.proteinatlas.org/). Enriched tissue and cell types were subsequently compared against the enriched tissues identified using LDSC analysis. This comparative approach was designed to explore how the genes associated with these phenotypes (i.e., WMHs, AD) relate to the cellular and tissue-specific enrichments found in the LDSC results. Interactions between significant WMH-and AD-associated genes were analysed using the STRING database (https://string-db.org/, accessed date: 19th Sep 2024). To identify enriched genetic pathways, k-means clustering (k=5) was applied to the interaction data obtained from STRING. Genes for WMHs were further compared to assess overlap with genes associated with AD (http://www.alzgene.org/).

### 2.5 Gene-set enrichment analysis

We used MAGMA (version 1.10) to identify significant genes sets associated with our phenotypes – WMHs and AD. Specifically, we used MAGMA gene-set analysis to assess over-representation of biological functions based on gene annotations using curated gene sets and gene ontology (GO) terms obtained from the Molecular Signature Database (MsigDB v5.2). In this context, over-representation refers to the comparison between the observed gene sets and the set of all genes considered in the GWAS analysis, including those mapped from SNPs during the gene analysis. Gene sets with Bonferroni-corrected *p* < 0.05 were considered to be significantly enriched.

## 3. RESULTS

### 3.1 Publicly Available GWAS Summary Statistics

Following our review of the PubMed literature and GWAS database (approach described in section 2.1), we identified three WMH GWAS studies with publicly available summary statistics ^19,20,38^ and sample sizes ranging from 11,226 to 33,200 participants. To ensure consistency in linkage disequilibrium patterns for LDSC analysis, following our review of GWAS catalogue for AD we included six studies published 2013 onwards due to their larger sample sizes (N = 74,046 to 636,674) and use of predominantly European cohorts ^39–44^. Further details on these studies for WMHs and AD are illustrated in Figure 1, which also provides a comprehensive summary of the sample sizes used.

**Figure 1:**
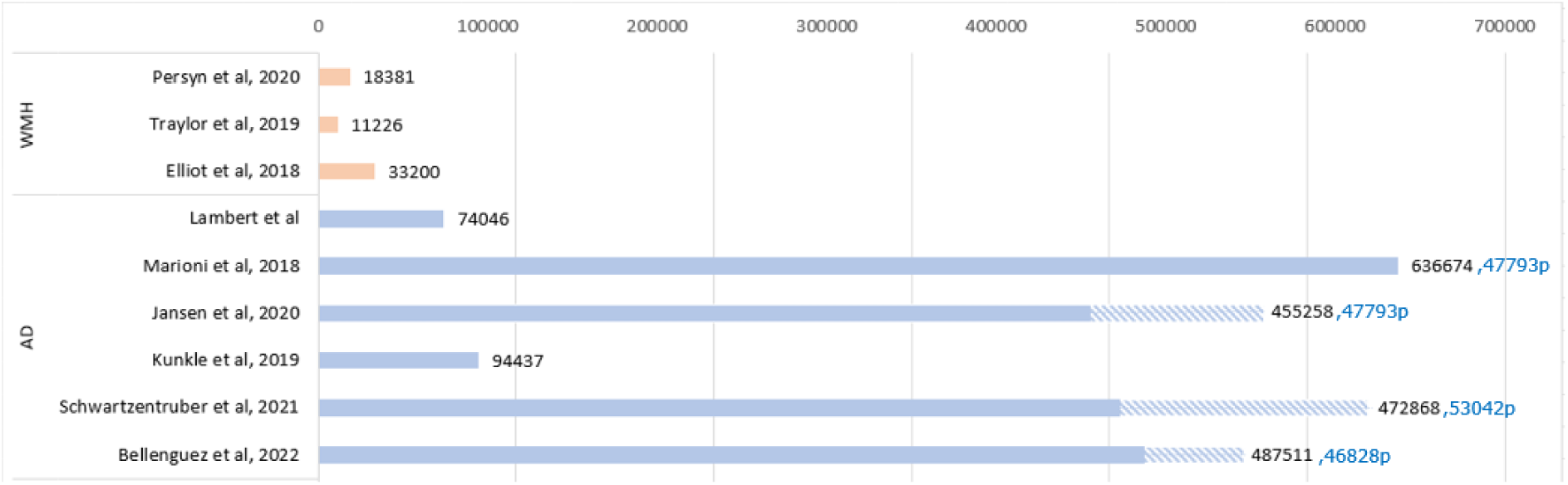
Bar plot showing number of participants per GWAS study included in our analyses. (“p” and diagonal line pattern indicates proxy cases). *Abbreviations:* AD, Alzheimer’s disease; WMH, White matter hyperintensity.

Although the study by Smith et al. ^38^ did not focus solely on WMHs, it remains relevant due to its use of the image-derived phenotype (IDP_T2_FLAIR_BIANCA_WMH_volume) for accurately quantifying WMH volumes. This phenotype, captured via the widely used automated BIANCA method ^45^ for WMH detection, enriched our analysis by increasing sample size and enhancing our understanding of the genetic basis of WMHs.

### 3.2 Tissue and Cell-Specific Partitioned Heritability Analysis Results

Following qualitative and statistical analyses, we identified a significant enrichment of WMH-associated SNPs in four distinct tissues. Displayed in Figure 2A, these findings were supported by both visual (greyed cells) and statistical indicators (greyed cells with an asterisk). WMH-associated-SNPs were significantly enriched in four tissues: Cardiovascular and kidney enrichments were observed in WMH only, while central nervous system (CNS) and liver enrichments were found common to both WMH-and AD-associated-SNPs (Figure 2A). Notably, we observed that the same three tissue types, i.e., immune, liver, and other were enriched in non-proxy AD studies (Figure 2A) as compared to CNS, connective bone, gastrointestinal, immune, liver, other for proxy AD studies (Figure 2A).

**Figure 2:**
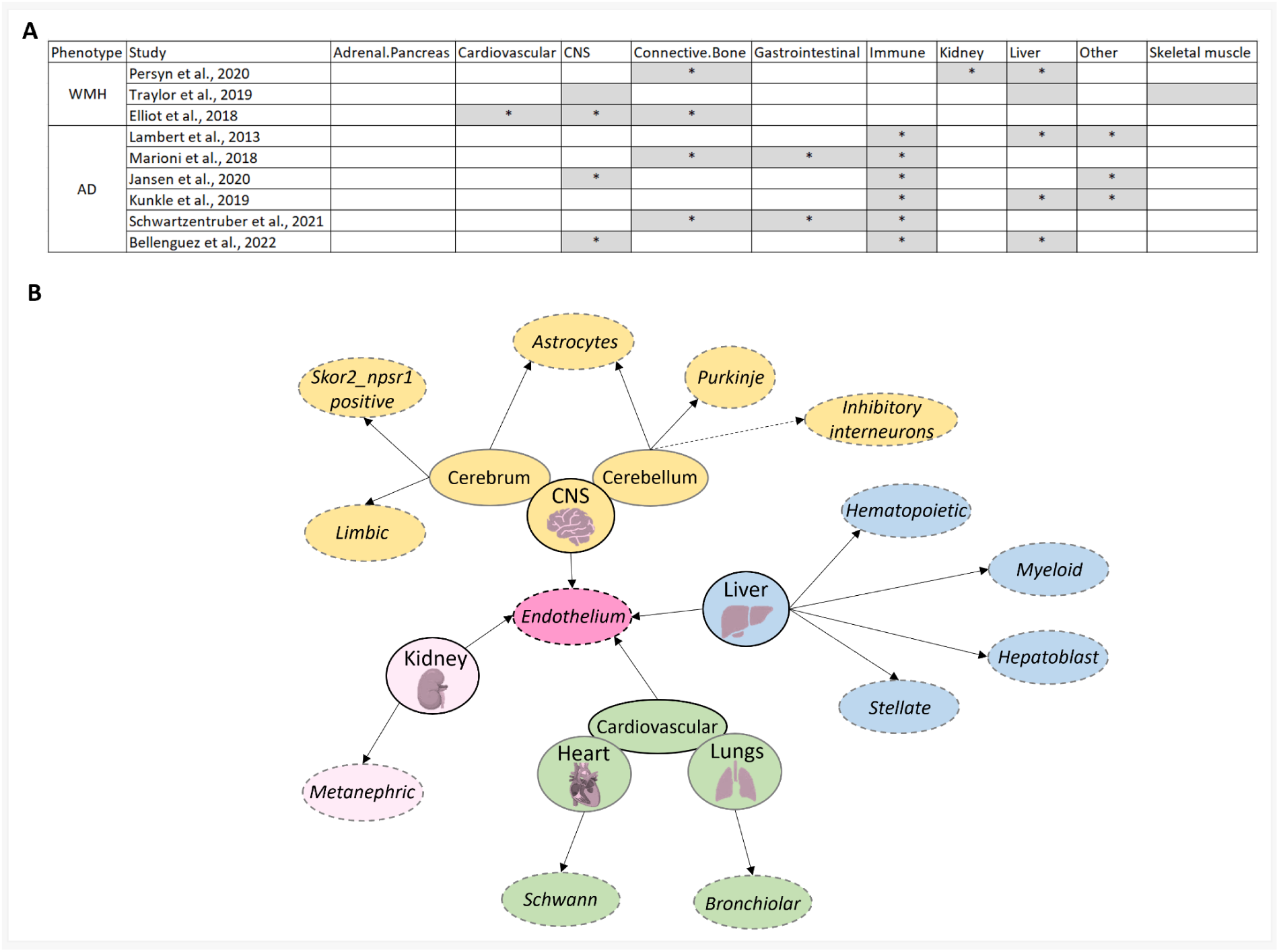
Enrichments using tissue and cell specific annotations for WMHs and AD. **A)** Enriched tissues for phenotypes (WMHs and AD) following false discovery rate correction which was applied on -log10(enrichement_p) values. * indicates tissues that survive false discovery rate correction. **B)** Graph showing cells (circle outlined using dashed lines) and tissue types (circle outlined using solid line) enriched for WMH associated-SNPs (solid arrow line) and cell-types enriched for overlapping tissues between WMHs and AD (dashed arrow line). *Abbreviations:* AD, Alzheimer’s disease; CNS, Central nervous system; WMH, White matter hyperintensity

Cell analysis within the four tissues enriched with WMH-associated-SNPs shows that 16/64 cell-types were also enriched, with vascular endothelial cells (vECs) being enriched in all four tissues. The tissue with the highest proportion of cell-types showing WMH-associated-SNPs enrichment was the liver (5/9 cell-types, 55%), followed by CNS (6/18 cell-types, 33%). While WMHs and AD both showed enrichment in CNS and liver tissues, in the CNS, cell-specific analysis highlighted enrichment in distinct cerebellar cell-types, with inhibitory interneurons and Purkinje cells being enriched for AD and WMHs, respectively (Figure 2B). Cell-specific analyses on liver cells showed no AD-associated-SNPs enrichment.

### 3.3 Gene-based Analysis Results

#### 3.3.1 MAGMA analysis findings

##### WMH findings

MAGMA gene analysis identified 39 genes significantly (*p* < 0.05/total number of protein-coding genes) associated with the WMH phenotype, across studies (N=3). Among these, four genes were recurrently associated in more than one study. Specifically, *MTHFD1L* was significantly associated with WMH phenotype in all three studies ^19,20,38^, while *NBR1*, *TMEM106A*, and *BRCA1* were found to be significant in two studies ^20,38^. Expression of these 39 genes in tissue types across the brain-body axis checked using human protein atlas showed high tissue specificity and enrichment for 18/39 genes with top five enriched tissues being: brain (8/39), liver (5/39), testis (4/39), kidney (3/39), and heart (3/39) (Table 1, Figure 3).

**Figure 3:**
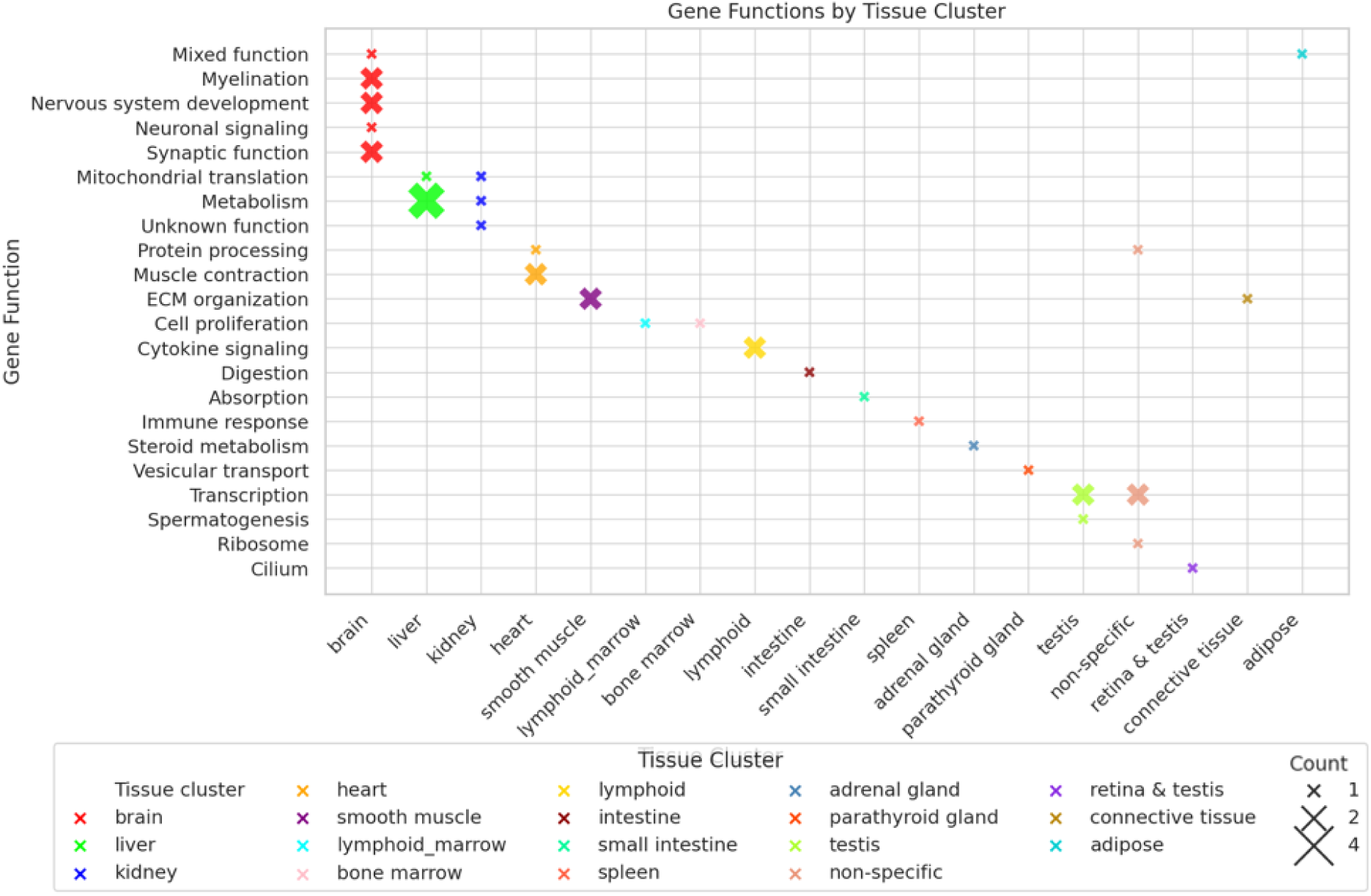
Plot showcasing the gene count for statistically significant genes associated with WMHs (found using MAGMA) expressed in different tissue types. Size of (x) represents the gene count.

**Table 1:** Tissue specificity of WMH associated genes (18/39) across the brain-body using human protein atlas (HPA). The 39 genes were found to be statistically significantly associated with WMHs in MAGMA gene analysis. The **bolded** gene was significantly associated with WMHs across all three studies, while the <u>underlined</u> genes were found to be significantly associated with WMHs in two studies. Clusters marked with ^ were found to be enriched for AD associated genes from MAGMA analysis. *Abbreviations:* ECM, Extracellular matrix; HPA, Human protein atlas.

| Gene symbol | Gene name (HPA) | Tissue (HPA cluster #) | Gene function (HPA) |
| --- | --- | --- | --- |
| *FAM167A* | Family with sequence similarity 167 member A | ^brain (#61) | Neuronal signalling |
| *MCF2L2* | MCF.2 cell line derived transforming sequence-like 2 | brain_neuronal (#17) | Synaptic function |
| *MTMR9* | Myotubularin related protein 9 | brain_neuronal (#17) | Synaptic function |
| *MTHFD1L* | Methylenetetrahydrofolate dehydrogenase (NADP+ dependent) 1 like | ^brain_cerebellum (#65) | Nervous system development |
| *PRAG1* | PEAK1 related, kinase-activating pseudokinase 1 | ^brain_cerebellum (#65) | Nervous system development |
| *KLHL32* | Kelch like family member 32 | brain_oligodendrocytes_ (#9) | Myelination |
| *RND2* | Rho family GTPase 2 | brain_oligodendrocytes_ (#9) | Myelination |
| *CCDC85A* | Coiled-coil domain containing 85A | ^brain_non-specific (#34) | Mixed function |
| *ARL5B* | ADP ribosylation factor like GTPase 5B | ^liver (#83) | Metabolism |
| *NSUN6* | NOP2/Sun RNA methyltransferase 6 | ^liver (#83) | Metabolism |
| *PPP1R3B* | Protein phosphatase 1 regulatory subunit 3B | ^liver (#83) | Metabolism |
| *G6PC1* | Glucose-6-phosphatase catalytic subunit 1 | ^liver & kidney (#1) | Metabolism |
| *MSRA* | Methionine sulfoxide reductase A | ^liver_kidney_non-specific (#43) | Mitochondrial translation |
| *TMEM106A* | Transmembrane protein 106A | ^kidney_thyroid gland (#51) | Unknown function |
| *GATA4* | GATA binding protein 4 | ^heart_non-specific (#68) | Protein processing |
| *PTGES3L* | Prostaglandin E synthase 3 like | ^heart_skeletal muscle (#14) | Muscle contraction |
| *PTGES3L-AARS1* | PTGES3L-AARS1 readthrough | ^heart_skeletal muscle (#14) | Muscle contraction |
| *VAT1* | Vesicle amine transport 1 | ^smooth muscle (#41) | ECM organisation |
| *PPP1R12A* | Protein phosphatase 1 regulatory subunit 12A | ^smooth muscle (#41) | ECM organisation |
| *BRCA1* | BRCA1 DNA repair associated | ^lymphoid tissue & bone marrow (#67) | Cell proliferation |
| ***DHX8*** | DEAH-box helicase 8 | ^bone marrow (#77) | Cell proliferation |
| *CTLA4* | Cytotoxic T-lymphocyte associated protein 4 | ^lymphoid (#36) | Cytokine signalling |
| *CD28* | CD28 molecule | ^lymphoid (#36) | Cytokine signalling |
| *CLDN23* | Claudin 23 | ^intestine (#45) | Digestion |
| *MFHAS1* | Multifunctional RCO family signalling regulator 1 | ^small intestine (#25) | Absorption |
| *IFI35* | Interferon induced protein 35 | ^spleen (#47) | Immune response |
| *SLC18A1* | Solute carrier family 18 member A1 | ^adrenal gland (#26) | Steroid metabolism |
| *ETV4* | ETS variant transcription factor 4 | ^parathyroid gland (#24) | Vesicular transport |
| *XKR6* | XK related 6 | testis (#46) | Spermatogenesis |
| *AARS1* | Alanyl-tRNA synthetase domain containing 1 | ^testis (#78) | Transcription |
| *NBR1* | NBR1 autophagy cargo receptor | ^testis (#78) | Transcription |
| *PILRB* | Paired immunoglobulin like type 2 receptor beta | ^testis (#78) | Transcription |
| *RPL27* | Ribosomal protein L27 | ^non-specific (#18) | Ribosome |
| ***RUNDC1*** | RUN domain containing 1 | ^non-specific (#66) | Transcription |
| *ERI1* | Exoribonuclease 1 | ^non-specific (#40) | Transcription |
| *TRIP11* | Thyroid hormone receptor interactor 11 | ^non-specific (#68) | Protein processing |
| *TNKS* | Tankyrase | ^retina & testis (#39) | Cilium |
| *LAMA4* | Laminin subunit alpha 4 | ^connective tissue (#62) | ECM organisation |
| *CTSB* | Cathepsin B | ^adipose (#7) | Mixed function |

##### AD findings

In our MAGMA gene analysis for AD, a total of 291 genes were found to be statistically significantly associated with AD (Supplementary Table S2) with 117/291 of them being associated significantly in more than one study. Expression of these 291 genes in tissue types across the brain-body axis checked using human protein atlas showed high tissue specificity and enrichment for 164/291 genes with top five enriched tissues being: brain (37/291, 12.7%), testis (30/291, 10.3%), liver (28/291, 9.6%), lymphoid (25/291, 8.6%), and bone marrow (17/291, 5.8%) (Supplementary Table S2). These findings are in-line with our findings from LDSC analysis where AD-associated SNPs were enriched not just in the CNS but also in the liver.

We observed overlap between genes associated with WMHs (after MAGMA analysis) and genes associated with AD (from Alzgene), specifically *ARL5B* and *MTHFD1L.* However we did not see any overlap between statistically significant MAGMA genes associated with WMHs and AD.

#### 3.3.2 Characterization of MAGMA-prioritised genes

Additional STRING network analyses of WMHs and AD associated genes (after MAGMA analyses) identified 15/39 WMH genes interacting with 60 AD associated genes (Table 2, Figure 4). Further k-means clustering analyses (k=5) of this interaction revealed pathways associated with immune response (cluster 1), DNA repair (cluster 2), spliceosome and ribosome biogenesis (cluster 3), autophagosome-lysosome fusion (cluster 4), and heart development (cluster 5) (Figure 4).

**Figure 4:**
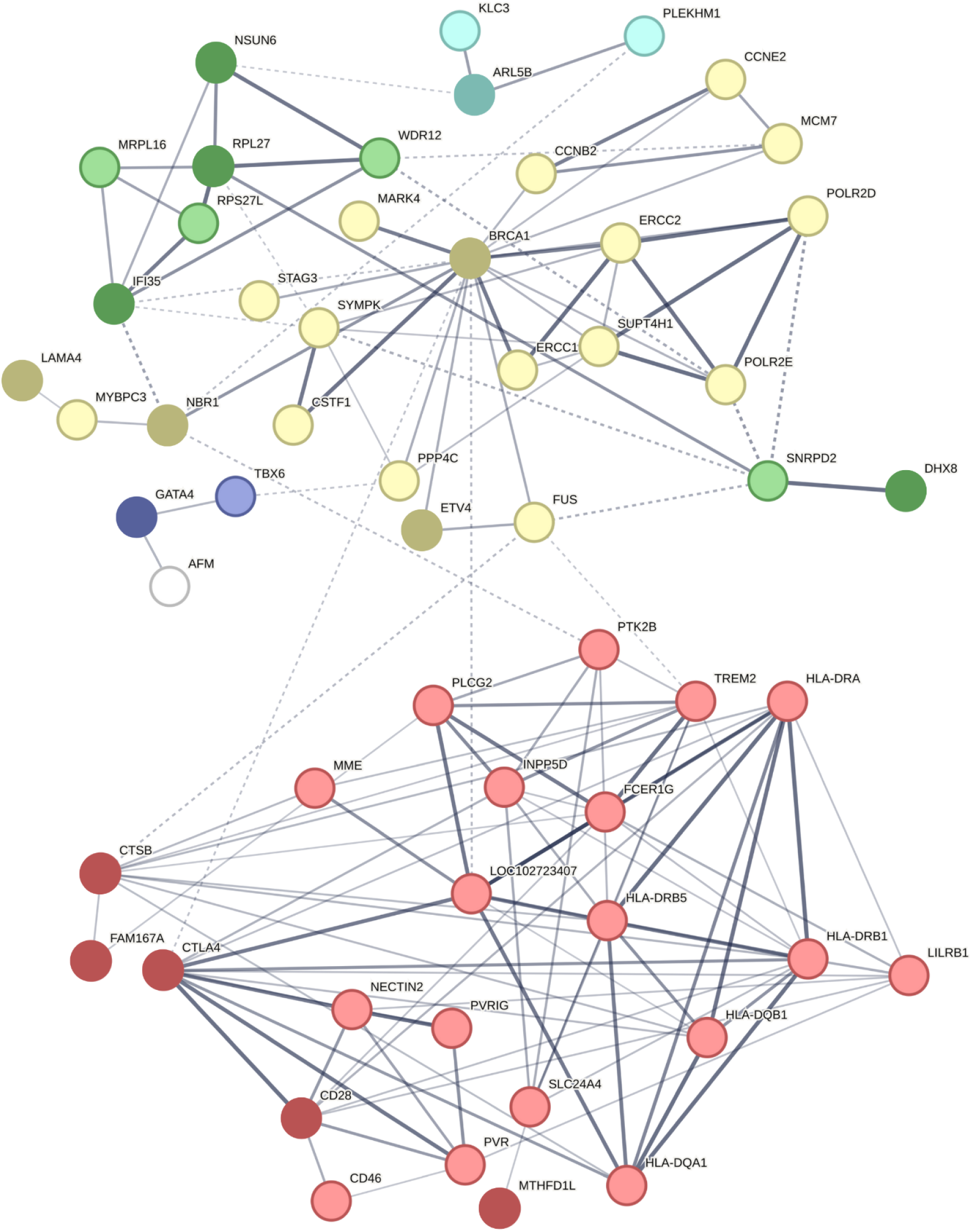
k-means cluster analyses (k=5) of interactions between statistically significant WMH and AD associated genes after MAGMA analyses (Confidence interval = 0.4). Dark shaded nodes represent WMH associated genes and light shaded nodes represent AD associated genes. Edges represent gene-gene associations and edge thickness (dotted to solid) represents the confidence of this association. Cluster 1 (red): immune response; Cluster 2 (yellow): DNA repair; Cluster 3 (green): spliceosome and ribosome biogenesis; Cluster 4 (blue): Autophagosome-lysosome fusion; Cluster 5 (purple): somitogenesis and heart development.

**Table 2:**
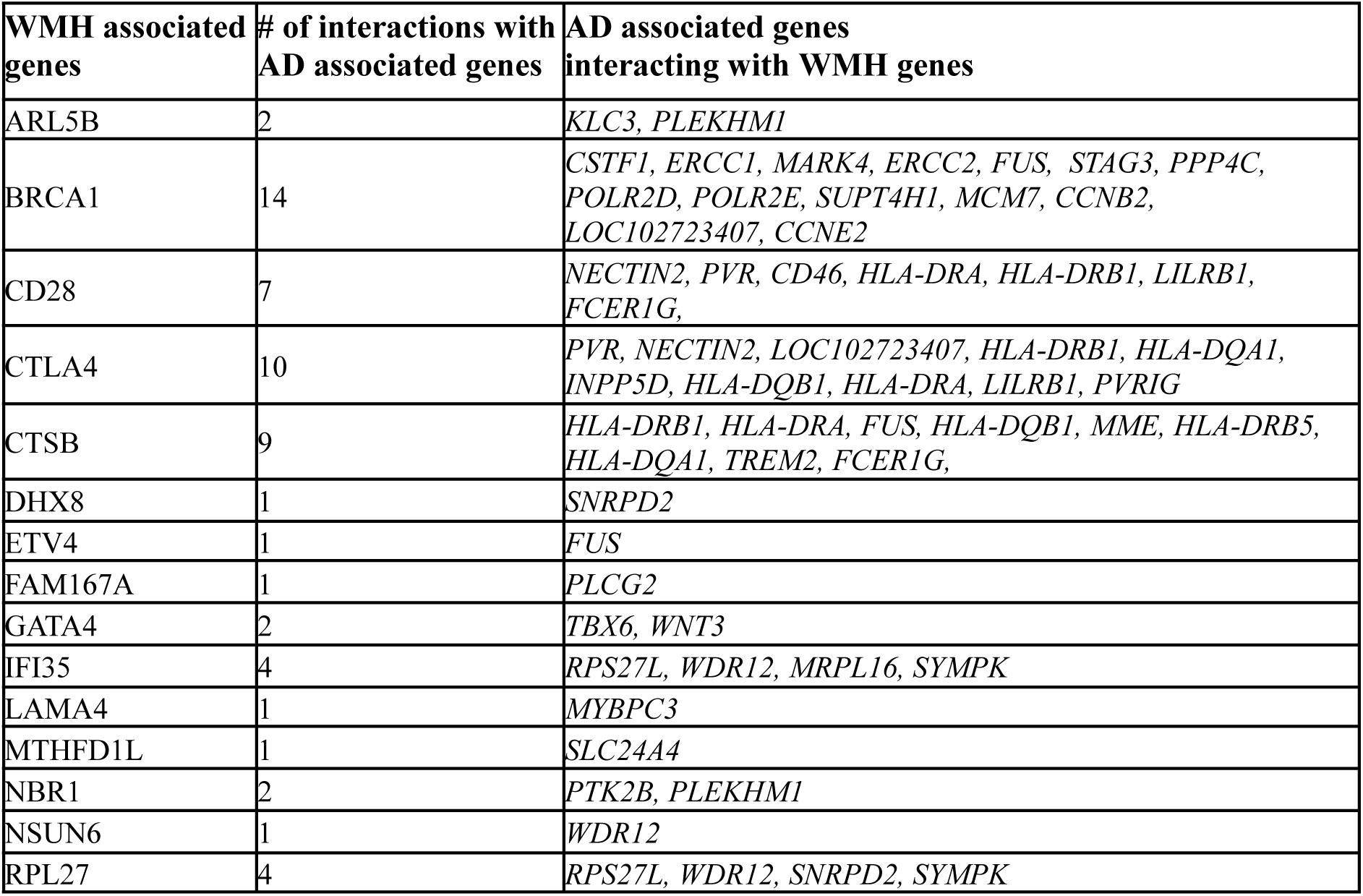
Interactions between statistically significant WMH and AD associated genes after MAGMA analyses using STRING.

| WMH associated genes | # of interactions with AD associated genes | AD associated genes interacting with WMH genes |
| --- | --- | --- |
| ARL5B | 2 | *KLC3, PLEKHM1* |
| BRCA1 | 14 | *CSTF1, ERCC1, MARK4, ERCC2, FUS, STAG3, PPP4C, POLR2D, POLR2E, SUPT4H1, MCM7, CCNB2, LOC102723407, CCNE2* |
| CD28 | 7 | *NECTIN2, PVR, CD46, HLA-DRA, HLA-DRB1, LILRB1, FCER1G,* |
| CTLA4 | 10 | *PVR, NECTIN2, LOC102723407, HLA-DRB1, HLA-DQA1, INPP5D, HLA-DQB1, HLA-DRA, LILRB1, PVRIG* |
| CTSB | 9 | *HLA-DRB1, HLA-DRA, FUS, HLA-DQB1, MME, HLA-DRB5, HLA-DQA1, TREM2, FCER1G,* |
| DHX8 | 1 | *SNRPD2* |
| ETV4 | 1 | *FUS* |
| FAM167A | 1 | *PLCG2* |
| GATA4 | 2 | *TBX6, WNT3* |
| IFI35 | 4 | *RPS27L, WDR12, MRPL16, SYMPK* |
| LAMA4 | 1 | *MYBPC3* |
| MTHFD1L | 1 | *SLC24A4* |
| NBR1 | 2 | *PTK2B, PLEKHM1* |
| NSUN6 | 1 | *WDR12* |
| RPL27 | 4 | *RPS27L, WDR12, SNRPD2, SYMPK* |

### 3.4 Gene-set enrichment Analysis Results

Our MAGMA gene set analysis for WMHs did not reveal any statistically significant genetic pathways while the analysis for AD highlighted multiple significant pathways (Supplementary Table S3).

## 4. DISCUSSION

Our study offers new insight into tissue-and cell-specific genetic enrichment patterns across the brain-body axis specific to WMHs, and common between WMHs and AD. Our tissue-specific analysis revealed that WMH-associated SNPs were significantly enriched in tissues beyond the CNS, namely liver, cardiovascular, and kidney – with liver being a common tissue enriched for both WMHs and AD. Our cell-specific analysis showed enrichment of vascular endothelial cells across the tissuel types enriched for WMHs, highlighting their central role in the development of WMHs. Additionally, our gene-level analysis highlighted overlapping patterns of tissue enrichment for both WMHs and AD, and showed interactions between WMH and AD associated genes.

### 4.1 Genetic Enrichment of WMH-associated SNPs in Peripheral Tissues

Since we found that WMH-associated SNPs were significantly enriched in liver, cardiovascular system, and kidney, their link with vascular brain injury has been discussed in Sections 4.1.1 to 4.1.3, respectively.

#### 4.1.1 Link between liver health and vascular brain injury

Using cross-sectional data from middle-aged to older adults (N=1,260), Jang et al. ^28^ reported that fatty liver disease (non-alcoholic variant) was significantly associated with moderate to severe cerebral WMH volumes, though not lacunes and microbleeds. This finding was reinforced by a longitudinal study (N=1,706) which showed that midlife non-alcoholic fatty liver disease predicted greater late-life WMH volume and a faster rate of WMH progression over six years ^46^. Additionally, midlife non-alcoholic fatty liver disease was linked to Alzheimer’s disease biomarkers, including lower plasma Aβ42:40 ratios and temporal-parietal cortical thinning ^46^. Systemic inflammation and vascular injury, including carotid atherosclerosis ^28,47–49^, are believed to underlie the liver-brain connection by weakening the blood-brain barrier and causing endothelial dysfunction, ultimately contributing to the development of WMHs ^50,51^. The proposed inflammation-based mechanism has since been supported by large-scale whole body imaging data from the UK Biobank (N > 30K), which demonstrated that liver inflammation and fibrosis were significantly associated with larger WMH volumes and perturbed white matter microstructure^52^. Reinforcing the vascular-based mechanism, the adiponutrin gene (*PNPLA3* [patatin-like phospholipase 3]; rs738409 [C > G]), a well-established non-alcoholic fatty liver disease risk variant, has been not only (a) associated with increased WMH volume and microbleeds ^53^, but also (b) linked to carotid atherosclerosis in select studies ^54^. Additionally, animal studies indicate that hyperammonemia associated with chronic liver disease, reduces cortical oxygenation, suggesting that impaired cerebral blood flow and oxygen supply may contribute to WMH development ^55^.

#### 4.1.2 Link between cardiovascular health and vascular brain injury

A review of studies (1973-2018) found that heart hypoperfusion, stemming from pathologies like cardiac small vessel disease, can contribute to WMHs by reducing cerebral blood flow ^56^. Supporting this, Mazini et al. ^57^ showed that in individuals with cardiac small vessel disease, higher resting left-ventricle myocardial blood flow correlated with increased deep grey matter WMHs. In hypertensive individuals, higher aortic pulse wave velocity, a marker of aortic stiffness, was significantly associated with greater whole-brain WMH volumes ^58^. Similarly, higher pulse pressure in hypertensive individuals was associated with WMHs. ^59^. A large body of literature indicates that the link between cardiac dysfunction and white matter hyperintensities is due to mechanisms such endothelial dysfunction, vascular ageing and fibrosis ^56^.

#### 4.1.3 Link between kidney health and vascular brain injury

Supporting our findings, emerging literature links kidney health to vascular brain injury. In particular, a significant association between decreased kidney function and a higher prevalence of WMHs was reported by Makin et al. ^27^. The authors suggested that factors such as hypertension may cause injury to both the kidney and the cerebrovasculature ^60,61^. Further supporting the kidney-WMH connection, two systematic reviews examined the relationship between chronic kidney disease and cerebral WMHs and found that (a) individuals with chronic kidney disease had significantly higher WMHs compared to controls ^62^, and (b) quantitatively determined WMH volume increased significantly in chronic kidney disease patients with lowered glomerular filtration rate, increased creatinine clearance, and higher urine albumin-to-creatinine ratio ^63^. The kidney and vascular brain injury connection is further reinforced by the increased cerebral WMH volume and impaired cerebrovascular reactivity in hemodialysis patients, often in advanced stages of chronic kidney disease ^64^. Moreover, longitudinal data (6-month) in individuals with cerebral small vessel disease demonstrated that those with renal impairment had significantly higher WMH volume than those without, with the association persisting after adjusting for hypertension but not after adjusting for age ^65^.

### 4.2 Cell-specific Enrichment of WMH-associated SNPs

Given that our cell-specific analysis showed WMH-associated SNPs enriched in a) vascular endothelial cells across tissue types enriched for WMHs, and b) brain cell types we will discuss them next. However, it should be noted that literature on the remaining cell types in the brain (limbic, Skor2_npsr1 positive) and other organs and their association with WMHs were sparse, and recommend additional future investigations.

#### 4.2.1 Systemic endothelial cell-type enrichment for WMH-associated SNPs

We demonstrated that SNPs associated with WMHs were enriched in vECs not only in the central nervous system, but also in cardiovascular (heart, lungs), liver, and kidney tissues. Our findings are supported by literature linking WMHs with systemic endothelial cells. Specifically, in older adults with cardiovascular disease, endothelial-dependent vasodilation has been reported to be inversely associated with WMH volumes ^66^. Similarly, in acute ischemic stroke patients elevated endothelial dysfunction markers (homocysteine and haemoglobin A1c ^67^) were independently associated with increased WMH volumes ^68^. In addition to the cardiovascular system, WMHs have also been associated with endothelial cells in the liver. Specifically, the liver plays a crucial role in regulating fibrinogen (coagulation marker), which has been linked to endothelial dysfunction and WMHs ^69,70^. Endothelium dysfunction in brain and kidneys have been shown (a) in low-grade chronic inflammation, which results in increased WMH volumes ^71^ and urine albumin excretion ^72,73^, and (b) with the association between albumin to creatinine ratio and WMH volumes ^73,74^.

#### 4.2.2 Brain cell-type enrichment for WMH-associated SNPs

We observed enrichment of WMH-associated SNPs in astrocytes within the cerebrum, which is consistent with literature implicating astrocytes in the development and progression of these lesions. Astrocytes play a crucial role in maintaining white matter integrity by (a) regulating ion-water homeostasis, preventing intra-myelin edema, and (b) supporting blood-brain barrier integrity. Astrocytic dysfunction can lead to impaired neurovascular coupling, increased susceptibility to ischemic damage, and a decline in blood-brain barrier structure, all of which are key factors in the development of WMHs ^75–78^. We also demonstrated different cerebellar cell types being enriched for WMHs (Purkinje cells). Substantial evidence also suggests that in AD neocerebellum, the activation of microglia and development of neurovascular inflammation are not associated with Purkinje cells count as measured through immunoreactivity to tau and ubiquitin proteins ^79^.

### 4.3 Gene level findings

Our gene-level analysis revealed certain overlapping patterns of tissue enrichment, and interactions between WMH-and AD-associated genes with newly identified WMH genes showing overlap with previously reported genes, including *KLHL32, TNKS,* and *XKR6* ^22^. We found several interactions between WMH and AD associated genes, in five clusters described below.

Immune activation (cluster 1) pathways are significant for both WMHs and AD, since white matter lesions and amyloid-beta plaques show elevated microglial activity, suggesting shared immune responses triggered by blood-brain barrier disruption. ^80–82^. Dysregulated DNA repair mechanisms (cluster 2) are implicated in oxidative DNA damage associated with white matter lesions, as studies show increased oxidative stress and impaired DNA repair in both WMHs and surrounding normal appearing white matter ^83,84^. Similarly, in AD, neurons exhibit heightened damage and reduced DNA repair capacity, contributing to progressive neurodegeneration ^85,86^. Dysregulated spliceosomal and ribosomal components (cluster 3) contribute to white matter damage, a hallmark of small vessel disease including the formation of WMHs. In CADASIL, a genetic form of cerebrovascular pathology, this pathway drives WMH development ^87^. Similarly, in AD, mislocalization of spliceosomal proteins to tau tangles disrupts normal cellular processes, contributing to neurodegeneration ^88^.

## 5. CONCLUSION

Overall, this study uncovers tissue-and cell-specific genetic enrichment patterns specific to WMHs and shared with AD. We unveiled that WMH-associated SNPs were enriched in tissues beyond the CNS, with liver being a common enriched tissue-type between WMHs and AD. Our analysis emphasised the central role of vascular endothelial cells in WMH development and also revealed interactions between genes associated with WMHs and AD.

Our MAGMA gene-analysis findings and subsequent analysis using the human protein atlas for WMHs and AD substantiated our s-LDSC findings, where WMH and AD-associated SNPs were enriched not just in the CNS but also in other tissue types (WMHs: liver, kidney, cardiovascular) (AD: liver, immune, gastrointestinal). Although our gene set analysis for WMHs did not reveal any statistically significant pathways due to the small sample size, a similar analysis highlighted well-known pathways implicated in AD.

Moreover, our findings provide new insights to the proposition that presence of MRI-detected WMHs is indicative of an underlying multi-system endothelial disorder affecting several vascular beds ^74,89^. We recommend further follow-up studies to enhance our understanding of the causative pathways associated with these multisystemic genetic findings.

## Data Availability

All data produced in the present work are contained in the manuscript

